# RoB2-Evaluator: Development and Technical Evaluation of a Rule-Constrained Large Language Model System for Cochrane Risk of Bias 2 Assessment

**DOI:** 10.64898/2026.09.22.26363693

**Authors:** Jithin Joseph, Rashmi Vishwanath, Jyothi Vettiyattusserril Sisirkumar

## Abstract

**Background:** Cochrane Risk of Bias 2 (RoB 2) assessment is methodologically demanding and resource intensive. Large language models may assist this process but can produce variable judgments.

**Aim:** To develop a rule-constrained LLM system for RoB 2 assessment and evaluate its temporal stability and agreement with human reviewers.

**Methods:** RoB2-Evaluator was developed using GPT-5.2 with fixed RoB 2 guidance and a locked instruction framework. Seven RCTs were used for calibration before version freezing. The frozen system was evaluated on 10 separate RCTs, with repeat assessment after one week and comparison against consensus judgments from two independent human reviewers.

**Results:** Temporal agreement was 90.0% (weighted κ = 0.93), and agreement with human consensus was also 90.0% (weighted κ = 0.93). Methodological rationales were stable in 94.3% of repeated comparisons. Full AI–human conceptual alignment occurred in 85.7%.

**Conclusion:** RoB2-Evaluator showed encouraging post-freeze stability, human agreement, and rationale consistency, supporting larger independent evaluations.

## Introduction

Risk-of-bias assessment is central to interpreting evidence from randomized control trials (RCTs) in systematic reviews. The Cochrane Risk of Bias 2 (RoB 2) tool provides structured signalling questions and decision algorithms across five core domains, with extensions for cluster-randomized and crossover trials, but its application remains methodologically demanding and resource intensive (1–3). Artificial Intelligence (AI) and Large language models (LLMs) may assist this process, yet previous evaluations have shown only modest-to-moderate agreement with human assessments and incomplete repeatability (4,5). We developed RoB2-Evaluator, a rule-constrained GPT-5.2 system using fixed RoB 2 guidance and locked decision instructions, and evaluated its temporal stability, agreement with human consensus judgments, and alignment of supporting methodological rationales.

## Methods

### Development and Calibration

RoB2-Evaluator was developed on GPT-5.2 using a fixed methodological knowledge base comprising the *Cochrane Handbook* Chapter 8, core RoB 2 guidance, and the cluster-randomized and crossover extensions (2,3). A locked instruction framework constrained assessments to the applicable RoB 2 signalling questions and decision pathways and restricted unsupported heuristics, numerical thresholds, and cross-domain inference. Web browsing and external database access were not used.

Development used seven purposively selected RCTs, three parallel-groups, two cluster-randomized, and two crossover trials. Phase 1 calibrated the five core RoB 2 domains, while Phase 2 calibrated the cluster and crossover extensions, with targeted refinement of recurrent decision drift. The underlying RoB 2 algorithms were not modified. Following calibration, the framework was frozen as RoB2-Evaluator Version 3. Characteristics of the development studies are provided in **Supplementary Table S1**, with detailed calibration procedures, operating constraints, and version control described in the Supplementary Methods. This methodological study used published trial reports and associated publicly available study documentation and did not involve recruitment of human participants or access to identifiable private data, hence ethics approval was not sought.

### Post-Freeze Evaluation

The frozen Version 3 system was evaluated using a separate set of 10 RCTs comprising four parallel-group, three cluster-randomized, and three crossover trials (**Supplementary Table S2**). No modifications to the instruction framework were permitted during evaluation. For each RCT, the full-text publication and any available protocol, statistical analysis plan, trial-registry record, and supplementary material were supplied. A standardized assessment prompt generated domain-level RoB 2 judgments, supporting methodological rationales, and an overall judgment. The default effect of interest was the effect of assignment to intervention. The complete assessment workflow, standardized prompts, and output procedures are provided in the **Supplementary Methods**.

### Temporal Stability

Temporal stability was assessed by repeating Version 3 evaluations of the 10 RCTs after one week in a new assessment session without access to the initial outputs, using the unchanged framework and assessment procedure.

### Agreement with Human Reviewers

Two reviewers with expertise in systematic review methodology and risk-of-bias assessment independently evaluated the same 10 RCTs using standard RoB 2 guidance and without access to RoB2-Evaluator outputs. Disagreements were resolved by consensus. RoB2-Evaluator judgments were compared with human consensus at domain and overall levels. One reviewer subsequently classified paired methodological rationales. Repeated AI rationales were categorized as identical, semantically equivalent, or different, while AI–human rationales were classified as full alignment, partial alignment, or conceptual disagreement using prespecified definitions detailed in the **Supplementary Methods**.

### Statistical Analysis

Temporal stability and AI–human agreement were assessed using percentage agreement, unweighted Cohen’s κ, and weighted Cohen’s κ to account for the ordinal structure of the RoB 2 categories. Ninety-five percent confidence intervals were reported for overall agreement coefficients. Extension-domain results were summarized descriptively because only three studies contributed to each extension. Rationale consistency was defined as the proportion of repeated methodological rationales classified as identical or semantically equivalent. Analyses were performed in R version 4.5.2 (R Foundation for Statistical Computing, Vienna, Austria).

## Results

### Development and Calibration

Calibration identified recurrent departures from intended RoB 2 decision pathways involving masking, open-label treatment, missing outcome data, recruitment in cluster-randomized trials, and carryover in crossover trials. Targeted instruction refinements were made without altering the underlying RoB 2 algorithms, after which the framework was frozen as RoB2-Evaluator Version 3. Detailed calibration findings are provided in the Supplementary Results a.

### Temporal Stability

Overall RoB 2 judgments were unchanged in 9 of 10 trials (90.0%), with unweighted Cohen’s κ = 0.85 (95% CI, 0.56–1.00) and weighted κ = 0.93 (95% CI, 0.64–1.00). Across the five core domains, agreement was 80.0% for D1 (κ = 0.60; 95% CI, 0.10–1.00), 60.0% for D2 (κ = −0.18; 95% CI, −0.46–0.11), 80.0% for D3 (κ = 0.41; 95% CI, −0.18–1.00), and 100.0% for both D4 and D5 (κ = 1.00; 95% CI, 1.00–1.00 for each). For the design-specific extensions, agreement was 100.0% for D1b and 66.7% for D6; κ estimates were not reported because only three trials contributed to each extension.

Across 53 paired domain-level methodological rationales, 31 (58.5%) were identical and 19 (35.8%) were semantically equivalent, yielding 94.3% rationale consistency. Three comparisons (5.7%) reflected substantively different rationales. All 53 rationales were logically consistent with their corresponding RoB 2 judgments. Detailed rationale comparisons are provided in **Supplementary Table S4**.

### Agreement with Human Reviewers

Overall RoB 2 judgments agreed with human consensus in 9 of 10 trials (90.0%), with unweighted Cohen’s κ = 0.85 (95% CI, 0.56–1.00) and weighted κ = 0.93 (95% CI, 0.64–1.00). Across the five core domains, agreement was 80.0% for D1 (κ = 0.60; 95% CI, 0.15–1.00), 90.0% for D2 (κ = 0.62; 95% CI, −0.04–1.00), 90.0% for D3 (κ = 0.74; 95% CI, 0.26–1.00), 100.0% for D4 (κ = 1.00; 95% CI, 1.00–1.00), and 70.0% for D5 (κ = 0.40; 95% CI, −0.05–0.85). Agreement for the design-specific extensions was 67.0% for D1b and 100.0% for D6; κ estimates were not reported because only three trials contributed to each extension.

Across 56 AI–human methodological-rationale comparisons, 48 (85.7%) showed full conceptual alignment and 8 (14.3%) showed partial alignment, with no conceptual disagreements. Domain-and study-level findings are provided in Supplementary Tables S5 and S6.

## Discussion

RoB2-Evaluator showed high overall temporal stability and agreement with human consensus after the assessment framework was frozen. Overall judgments were unchanged in 9 of 10 repeated assessments and agreed with human consensus in 9 of 10 studies; both comparisons yielded Cohen’s κ = 0.85 and weighted κ = 0.93. Methodological rationales were identical or semantically equivalent in 94.3% of repeated comparisons, while 85.7% of AI-human rationale comparisons showed full conceptual alignment and the remainder partial alignment, with no conceptual disagreements. These findings provide preliminary evidence that a rule-constrained LLM can apply RoB 2 with relatively consistent judgments and methodological rationales.

Temporal stability varied across domains, with the greatest variation for deviations from intended interventions (D2; 60% agreement; κ = −0.18). This contrasts with the higher overall stability and illustrates the importance of examining domain-level performance rather than relying solely on overall classifications. Rose et al. reported 74.7% agreement (κ = 0.62) between repeated ChatGPT-4o assessments using the earlier RoB1 framework, suggesting that repeatability remains an important consideration for LLM-assisted risk-of-bias assessment (4).

Previous evaluations of general-purpose LLMs have generally reported modest agreement with human risk-of-bias assessments (6–8). Taneri reported a weighted κ of 0.51 between ChatGPT-4o and Cochrane RoB 2 assessments across 84 RCTs (5). Agreement observed with RoB2-Evaluator was numerically higher, although direct comparison is limited by differences in models, trial samples, source materials, prompting, and evaluation design. One possible explanation is the use of fixed RoB 2 guidance, locked decision instructions, targeted correction of decision drift, and version freezing. Structured grounding has similarly been associated with improved LLM performance in other biomedical applications (9,10), although the present study cannot isolate the contribution of this architecture because no unconstrained GPT-5.2 comparator was included.

The potential value of standardization should also be considered alongside known variability in human risk-of-bias assessment. Inter-rater reliability for RoB 2 has been reported to be low overall, with substantial variation across domains (11), while earlier Cochrane studies similarly found marked inter-review variability (12,13). RoB2-Evaluator should therefore be viewed as a decision-support tool that may assist second-reviewer assessment, consistency checking, and adjudication rather than replace human methodological review.

Several limitations warrant emphasis. The post-freeze evaluation included only 10 purposively selected RCTs, with three studies contributing to each design-specific extension, limiting precision and generalizability. Domain-level κ estimates may be unstable because of the small sample and restricted category distribution. Rationale alignment was classified by a single reviewer, studies were assessed sequentially within a continuous workflow, and the underlying proprietary model may evolve independently of the locked framework. Efficiency and usability were not evaluated.

## Conclusion

RoB2-Evaluator demonstrated encouraging temporal stability, agreement with human consensus, and rationale consistency in this initial technical evaluation. Larger independent prospective studies, including direct LLM comparators and broader trial designs, are required before routine use.

## Generative AI Disclosure

RoB2-Evaluator, a custom GPT developed using GPT-5.2 (OpenAI), was used to generate the risk-of-bias assessments evaluated in this study. The system was configured with a fixed RoB 2– specific methodological knowledge base and locked instruction framework, as described in the Methods and Supplementary Methods. All outputs included in the analyses were evaluated according to the prespecified study procedures. ChatGPT was additionally used to assist with language editing and manuscript organization. All AI-assisted text was reviewed and verified by the authors, who take full responsibility for the final manuscript.

## Data and Materials Availability

Study-level data supporting the reported analyses are provided in the manuscript and Supplementary Material. The RoB2-Evaluator custom GPT is publicly accessible at https://chatgpt.com/g/g-699fa23342f881919cce20689276b1b3-rob2-evaluator. The Supplementary Material contains the operating instructions, standardized prompts, output templates, and version-control procedures used in the study.

## Supplementary Methods and Results

### Detailed Methods

#### Development of RoB2-Evaluator

RoB2-Evaluator was developed using GPT-5.2 as a rule-constrained large language model system for application of the Cochrane Risk of Bias 2 (RoB 2) tool to randomized controlled trials. The initial system incorporated a fixed methodological knowledge base comprising the *Cochrane Handbook* (Chapter 8), the core RoB 2 guidance, and the cluster-randomized and crossover extensions (1,2). A locked instruction framework constrained assessments to the RoB 2 signalling questions and decision algorithms and restricted unsupported heuristic rules, numeric thresholds, cross-domain inference, and use of information outside the uploaded trial documents and fixed methodological guidance. Web browsing and external database access were not used. This initial implementation was designated RoB2-Evaluator Version 1.

Development used seven purposively selected RCTs representing contrasting methodological scenarios: three parallel-groups, two cluster-randomized, and two crossover trials. These studies were used for calibration and correction of decision drift, not for estimation of system performance. Study characteristics are provided in **Supplementary Table S1**.

#### Phase 1: Parallel-group calibration and Version 2 freeze

Version 1 was calibrated using three parallel-group RCTs selected to challenge the five core RoB 2 domains across varied methodological scenarios. Recurrent decision drift was corrected through targeted instruction refinement, including inappropriate escalation related to open-label treatment, masking, missing-data thresholds, and cross-domain inference. The underlying RoB 2 algorithms were unchanged. After stable application of Domains 1–5, the framework was frozen as **RoB2-Evaluator Version 2**.

#### Phase 2: Extension-domain calibration and Version 3 freeze

Version 2 was calibrated using two cluster-randomized and two crossover trials selected to challenge the design-specific RoB 2 extensions. Cluster trials were used to test interpretation of recruitment timing, knowledge of cluster allocation, potential differential recruitment, and baseline imbalance. Crossover trials were used to test recognition of washout, persistence of intervention effects, treatment order, carryover plausibility, and analytical mitigation. Where assessments departed from the intended RoB 2 decision pathway, the instruction framework was refined without modification of the underlying RoB 2 algorithms. Following completion of extension-domain calibration, the framework was frozen as **RoB2-Evaluator Version 3** and used unchanged for subsequent post-freeze evaluation.

#### Post-freeze evaluation dataset

Ten separate RCTs comprised the post-freeze evaluation dataset: four parallel-group, three cluster-randomized, and three crossover trials. The studies were purposively selected to represent heterogeneous methodological scenarios and were separate from those used during development and calibration. Characteristics are provided in **Supplementary Table S2**

#### Temporal stability

Temporal stability was evaluated using Version 3 across the 10 evaluation RCTs at two assessment occasions (T1 and T2), separated by one week. The instruction framework, methodological guidance, and assessment workflow remained unchanged. Domain-level and overall RoB 2 judgments and their accompanying methodological rationales were compared between assessments.

#### Agreement with human reviewers

Two reviewers with expertise in systematic review methodology and risk-of-bias assessment independently performed RoB 2 assessments according to standard Cochrane guidance, without access to RoB2-Evaluator outputs. Disagreements were resolved through consensus, and the resulting consensus judgments served as the human comparator. RoB2-Evaluator Version 3 outputs were compared with human consensus at the domain and overall levels. AI–human methodological rationales were subsequently compared by one reviewer and classified as full alignment, partial alignment, or conceptual disagreement according to prespecified definitions.

#### RoB2-Evaluator assessment workflow

For each RCT, the evaluator uploaded the full-text publication together with any available protocol, statistical analysis plan, trial-registry record, and supplementary material. A standardized assessment prompt initiated the RoB 2 evaluation, producing domain-specific reasoning, domain-level judgments, and an overall risk-of-bias classification. Studies were assessed sequentially within the same assessment workflow until all included RCTs had been completed. After the final study, a standardized output prompt generated two consolidated outputs: a RoB2 Summary Table containing study-level domain and overall judgments and a RoB2 Master Table containing each domain judgment together with its supporting methodological rationale. The present study evaluated the assessment process rather than automated generation of publication-ready visualizations. The standardized prompts, output templates, operating constraints, and table-generation procedures are reproduced in **Supplementary Appendix 1**.

#### Statistical analysis

Temporal stability and AI–human agreement were quantified using percentage agreement, unweighted Cohen’s κ, and weighted Cohen’s κ to account for the ordinal structure of the RoB 2 categories (*Low risk*, *Some concerns*, and *High risk*). Ninety-five percent confidence intervals were calculated for agreement coefficients. Extension-domain results were summarized descriptively because only three studies contributed to each extension. Temporal methodological rationales were classified as identical, semantically equivalent, or different; rationale consistency was defined as the proportion classified as identical or semantically equivalent. Reason–decision alignment assessed whether each methodological rationale logically supported its corresponding RoB 2 judgment. AI–human methodological rationales were classified as full alignment, partial alignment, or conceptual disagreement according to prespecified definitions. Analyses were performed in R version 4.5.2 (R Foundation for Statistical Computing, Vienna, Austria).

### Supplementary Results

#### Core-Domain Calibration

Core-domain calibration identified recurrent departures from intended RoB 2 logic related to heuristic interpretation of open-label interventions, masking, missing outcome data, and domain-specific methodological information. Targeted refinements prevented automatic escalation based on lack of masking alone, prohibited unsupported numerical thresholds for missingness, reinforced the distinction between allocation concealment and blinding, and strengthened domain independence. These changes clarified implementation of the existing RoB 2 decision pathways without modifying the underlying algorithms. Following stable application of Domains 1–5, the framework was frozen as **Version 2**.

#### Extension-domain calibration

In cluster trials, early assessments tended to over-escalate recruitment bias when participants were enrolled after cluster randomization. Peiris et al. (2015) helped identify this problem, and the instructions were refined so that post-randomization recruitment was interpreted together with knowledge of cluster allocation, the potential for differential recruitment, and supporting evidence such as baseline imbalance, rather than being treated as sufficient on its own to determine Domain 1b (3). Taneja et al., which reported a 4-day washout and assessment of treatment order/carryover, was used to reinforce that crossover design alone should not trigger a high-risk judgment (4). Holze et al., where persistent treatment effects made carryover clinically important and first-period analyses were emphasized, was used to strengthen recognition of situations in which carryover materially affects interpretation(5). After these refinements, the extension-domain framework was frozen as RoB2-Evaluator Version 3 and used unchanged for subsequent temporal-stability and human-agreement analyses.

#### Temporal Stability

RoB2-Evaluator showed high temporal stability across repeated assessments. Overall RoB 2 judgments were unchanged in 9 of 10 studies (90.0%), corresponding to an unweighted Cohen’s κ of 0.85 (95% CI, 0.56–1.00) and a weighted κ of 0.93 (95% CI, 0.64–1.00). Across the five core domains, agreement was 80.0% for D1 (κ = 0.60), 60.0% for D2 (κ = −0.18), 80.0% for D3 (κ = 0.41), and 100.0% for D4 and D5 (κ = 1.00 for both). Weighted and unweighted κ estimates were identical for D1–D5. For the design-specific extensions, agreement was 100.0% for D1b and 66.7% for D6; these results were interpreted descriptively because only three studies contributed to each extension. Summary estimates are presented in **Table 1 and Figure 1 of the main manuscript**.

**Figure 1.**
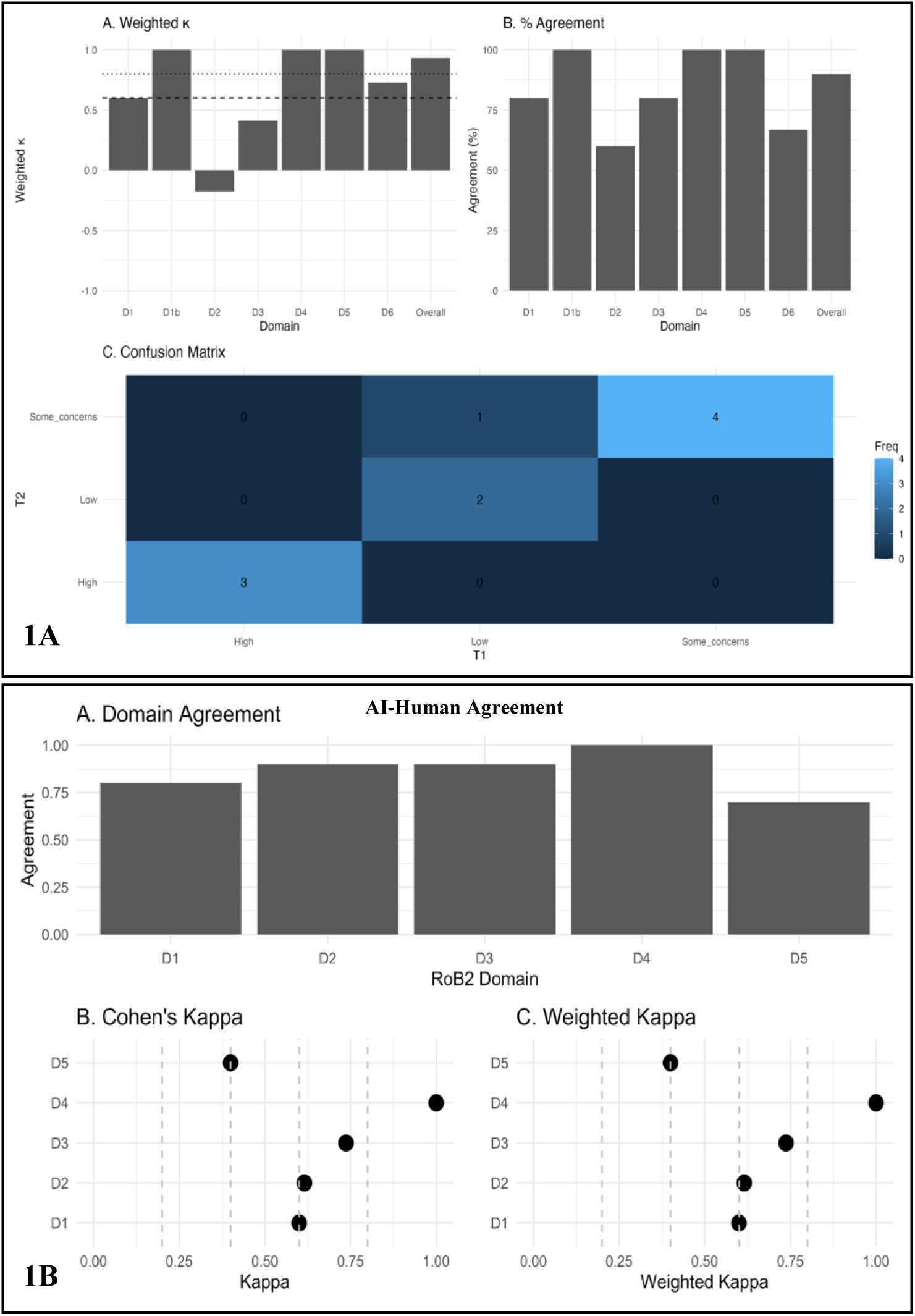
Post-freeze performance of RoB2-Evaluator. Upper panels (1A) show temporal stability using weighted κ, percentage agreement, and an overall-judgment confusion matrix. Lower panels (1B) show agreement with human consensus across core RoB 2 domains using percentage agreement, unweighted κ, and weighted κ.

**Table 1.** Temporal stability and agreement between RoB2-Evaluator and human consensus assessments.

| Assessment | n | Temporal agreement % | Temporal Cohen's $\kappa$ (95% CI) | AI-human agreement, % | AI-human Cohen's $\kappa$ (95% CI) |
| --- | --- | --- | --- | --- | --- |
| Overall RoB judgment | 10 | 90.0 | 0.85 (0.56–1.00);<br>weighted $\kappa$ = 0.93 (0.64–1.00) | 90.0 | 0.85 (0.56–1.00);<br>weighted $\kappa$ = 0.93 (0.64–1.00) |
| D1:Randomization process | 10 | 80.0 | 0.60 (0.10–1.00) | 80.0 | 0.60 (0.15–1.00) |
| D2: Deviations from intended interventions | 10 | 60.0 | –0.18 (–0.46–0.11) | 90.0 | 0.62 (–0.04–1.00) |
| D3: Missing outcome data | 10 | 80.0 | 0.41 (–0.18–1.00) | 90.0 | 0.74 (0.26–1.00) |
| D4: Measurement of the outcome | 10 | 100.0 | 1.00 (1.00–1.00) | 100.0 | 1.00 (1.00–1.00) |
| D5: Selection of the reported result | 10 | 100.0 | 1.00 (1.00–1.00) | 70.0 | 0.40 (–0.05–0.85) |
| D1b: Participant recruitment* | 3 | 100.0 | NA | 67.0 | NA |
| D6: Carryover effects† | 3 | 66.7 | NA | 100.0 | NA |

**Abbreviations:** CI, confidence interval; $\kappa$ , Cohen's kappa; RoB, risk of bias.
\*D1b was assessed only in cluster-randomized trials (n = 3).
† D6 was assessed only in crossover trials (n = 3). $\kappa$ estimates for extension domains were not reported because of the small number of applicable trials.
‡ Weighted and unweighted $\kappa$ estimates were identical for the five core domains (D1–D5); therefore, weighted $\kappa$ is reported separately only for the overall RoB judgment.

At the study level, the only change in overall RoB 2 classification occurred for Sha et al. (2015), which changed from *Low risk* at T1 to *Some concerns* at T2. Domain-level changes were more frequent but did not necessarily alter the overall judgment. Changes occurred in D1 for Sha et al. (2015) and Broughton et al. (1997); in D2 for Mukherjee et al. (2023), Ahmad et al. (2021), Garcia et al. (2020), and Cox et al. (2020); in D3 for Mukherjee et al. (2023) and Baeken et al. (2013); and in D6 for Sha et al. (2015). D1b remained unchanged in all applicable cluster-randomized trials. Detailed study-level findings are provided in **Supplementary Table S3**.

#### Methodological-Rationale Stability

Temporal consistency of the accompanying methodological rationales was evaluated across 53 paired domain-level assessments. Of these, 31 (58.5%) were classified as identical and 19 (35.8%) as semantically equivalent, yielding an overall rationale-consistency rate of 94.3% (50/53). Three comparisons (5.7%) reflected substantively different methodological rationales. Across all 53 comparisons, the generated rationale was logically consistent with its corresponding RoB 2 judgment, resulting in 100% reason–decision alignment. This measure reflects internal consistency between the stated rationale and assigned judgment and should not be interpreted as evidence of judgment accuracy. Definitions and complete rationale-consistency results are provided in **Supplementary Table S4**

#### Agreement with Human Reviewers

RoB2-Evaluator showed high agreement with human consensus assessments across the 10 post-freeze evaluation studies. Overall RoB 2 judgments agreed in 9 of 10 studies (90.0%), corresponding to an unweighted Cohen’s κ of 0.85 (95% CI, 0.56–1.00) and a weighted κ of 0.93 (95% CI, 0.64–1.00). Across the five core RoB 2 domains, agreement ranged from 70.0% to 100.0%. Agreement was 80.0% for D1 (κ = 0.60), 90.0% for D2 (κ = 0.62), 90.0% for D3 (κ = 0.74), 100.0% for D4 (κ = 1.00), and 70.0% for D5 (κ = 0.40). Weighted and unweighted κ estimates were identical for D1–D5. For the design-specific extensions, agreement was 67.0% for D1b and 100.0% for D6; these results were summarized descriptively because only three studies contributed to each extension. Summary agreement estimates are presented in **Table 1 and Figure 1 of the main manuscript**.

At the study level, disagreements were concentrated in a limited number of study-domain combinations. D1 disagreements occurred for Mukherjee et al. (2023) and Sha et al. (2015), while D5 disagreements occurred for Ahmad et al. (2021), Mukherjee et al. (2023), and Garcia et al. (2020). Single disagreements were observed for D2 in Garcia et al. (2020) and D3 in Hayes et al. (2019). Complete agreement was observed for D4 across all 10 studies. The study-level distribution of agreement and disagreement is shown in **Supplementary Figure S1**.

**Figure S1.**
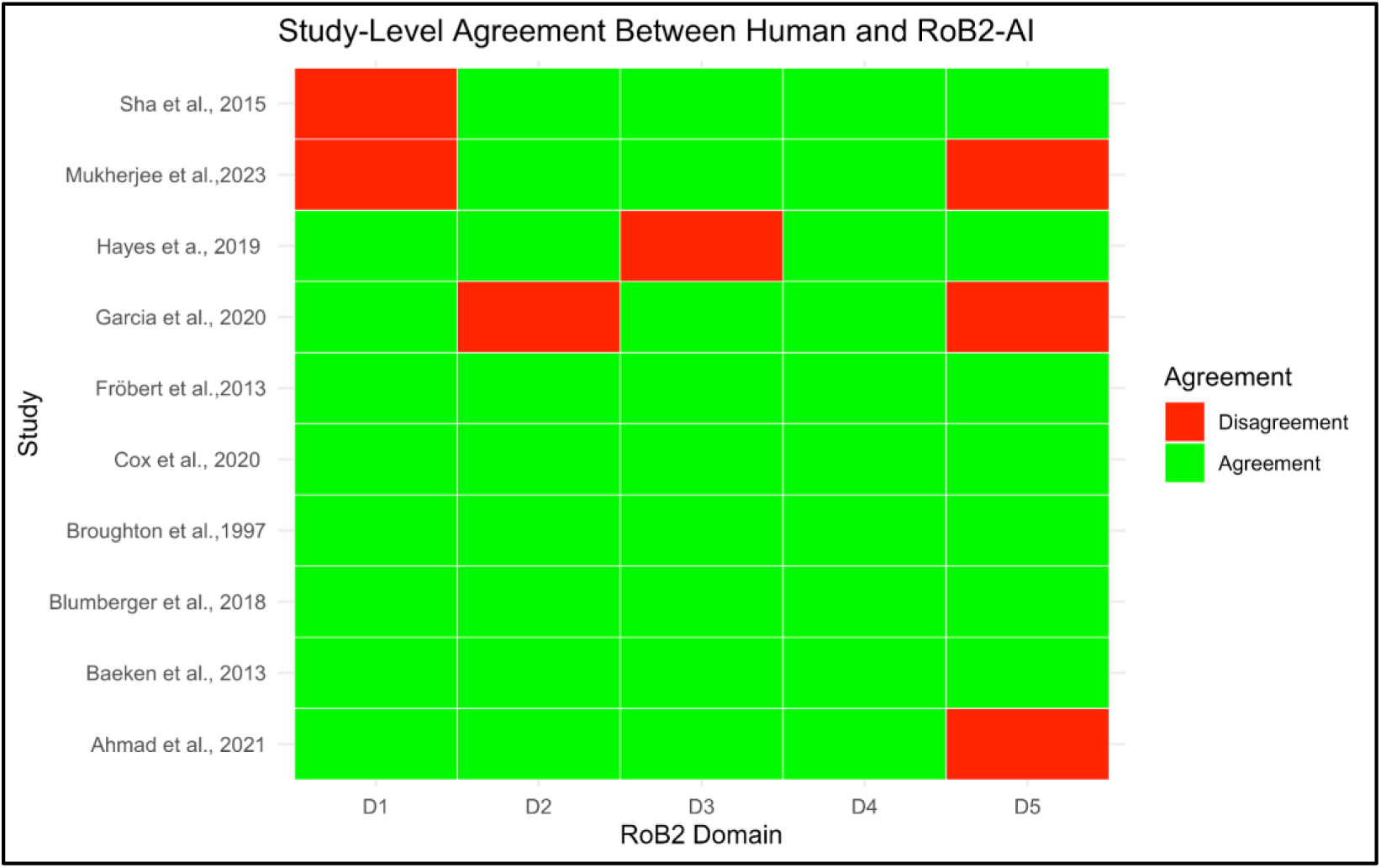
Study-level agreement between RoB2-Evaluator and human consensus assessments across the five core RoB 2 domains. Each cell represents agreement (green) or disagreement (red) between the AI-generated and human consensus risk-of-bias judgments for an individual study and RoB 2 domain. D1–D5 denote the five core domains of the Cochrane Risk of Bias 2 (RoB 2) tool.

#### Conceptual Alignment of AI-Generated Reasoning

Beyond agreement in RoB judgments, we evaluated whether RoB2-Evaluator identified the same underlying methodological issues as the human consensus assessment. Across 56 domain-level comparisons, 48 (85.7%) demonstrated full conceptual alignment and 8 (14.3%) demonstrated partial alignment, with no conceptual disagreements observed **(Supplementary Table S5)**. Full alignment was observed for all assessments of measurement of the outcome (D4), participant recruitment in cluster-randomized trials (D1b), and carryover effects in crossover trials (D6). Partial alignment occurred most frequently for the randomization process (D1) and selection of the reported result (D5). In these cases, RoB2-Evaluator and the human reviewers identified the same underlying methodological issue but differed in the detail or emphasis of the supporting rationale. Conceptual alignment assessed the methodological basis of the reasoning rather than concordance of the final RoB category; consequently, conceptual alignment could be present even when the resulting domain-level judgments differed. Representative study-level comparisons of AI- and human-generated reasoning are provided in **Supplementary Table S6**.

### Supplementary Discussion

The domain-level findings provide additional context beyond the overall agreement estimates reported in the main manuscript. Temporal agreement was lowest for deviations from intended interventions (D2), whereas AI–human agreement was lowest for selection of the reported result (D5). These domains require interpretation of contextual and analytical information rather than simple extraction of reported trial characteristics. D2 requires consideration of deviations, adherence, masking, and analytical handling in relation to the effect of assignment to intervention, while D5 may require comparison of publications with protocols, registries, or statistical analysis plans to determine whether a reported result was selected from multiple eligible outcomes or analyses (2). These findings support examination of domain-level behavior even when overall agreement is high.

Methodological-rationale analyses provided information not captured by categorical agreement. Across repeated assessments, 94.3% of rationales were identical or semantically equivalent, although three comparisons reflected substantively different methodological interpretations. Similarly, 85.7% of AI–human comparisons showed full conceptual alignment and the remainder partial alignment, with no conceptual disagreements. Conceptual alignment was evaluated independently of the final RoB 2 category; therefore, the same underlying methodological concern could be identified despite differences in how it was translated into a categorical judgment. The observed 100% reason–decision alignment should likewise be interpreted as internal logical consistency rather than evidence of judgment accuracy.

The design-specific extensions also highlighted methodological issues that may not be captured adequately by generic prompting. In cluster-randomized trials, calibration reinforced that recruitment after cluster randomization should not independently determine D1b; recruitment timing must be interpreted alongside knowledge of allocation, the potential for differential recruitment, and supporting evidence. In crossover trials, carryover assessment required consideration of intervention persistence, washout, treatment sequence, and analytical mitigation. Taneja et al. illustrated circumstances in which potential carryover was addressed through washout and analytical assessment, whereas Holze et al. illustrated situations in which persistent treatment effects made carryover clinically important (4,5). Because only three evaluation studies contributed to each extension, these findings remain exploratory.

Human consensus was used as the comparator but should not be interpreted as an error-free reference standard. Previous evaluations have documented substantial inter-rater variability in RoB assessment, including low overall reliability with RoB 2 and marked domain-level disagreement across Cochrane assessments (6–8). Rationale-level comparison was therefore useful for distinguishing categorical disagreement from fundamentally different methodological interpretations.

### Limitations

Several limitations require emphasis. The post-freeze evaluation included only 10 purposively selected RCTs, with three studies contributing to each design-specific extension, limiting precision and generalizability. The small sample and restricted distribution of RoB categories may also destabilize domain-level κ estimates. No direct comparison with unconstrained GPT-5.2 or another LLM was performed, so the independent contribution of the rule-constrained architecture cannot be determined. Temporal rationale consistency and AI–human conceptual alignment were classified by a single reviewer and may therefore be subject to interpretive bias. Studies were assessed sequentially within a continuous workflow, introducing possible context dependence across assessments. Finally, RoB2-Evaluator relies on a proprietary underlying model and execution environment that may evolve independently of the locked instruction framework. The study did not evaluate efficiency, usability, time savings, or downstream effects on systematic-review conclusions. Larger prospective evaluations using independent datasets, external reviewers, direct model comparators, and broader representation of cluster-randomized and crossover trials are therefore required.

---

**Supplementary Table S1.** Characteristics and methodological features of studies used during development and calibration of RoB2-Evaluator.

| Study ID | Trial design | Clinical context / intervention | Key methodological features represented | RoB 2 component / developmental purpose |
| --- | --- | --- | --- | --- |
| Allgulander et al. 2006 (9) | Parallel-group RCT | Escitalopram for generalized anxiety disorder | Open-label run-in followed by double-blind randomization; computer-generated sequence; identical medication; relapse outcome | Core domains; randomization, deviations, outcome measurement, and missing outcome data |
| Dobson et al. 2008 (10) | Parallel-group RCT | Behavioral activation, cognitive therapy, and antidepressant medication for major depression | Pharmacological and psychological interventions; differential masking feasibility; treatment withdrawal; relapse/recurrence outcomes | Core domains; particularly D2 and D4 in psychotherapy/open-label settings |
| Molenaar et al. 2020 (11) | Parallel-group RCT | Preventive cognitive therapy and antidepressant discontinuation during pregnancy | Stratified block randomization; nonpharmacological versus medication strategy; masked outcome assessors; small sample | Core domains; particularly D1–D4 |
| Peiris et al.<br>2015 (12) | Cluster-RCT | Cluster-level<br>healthcare<br>intervention | Participant recruitment relative to cluster<br>randomization; differential recruitment<br>and baseline imbalance considerations | Core domains and cluster<br>extension; recruitment-related bias (D1b) |
| Delgadillo et al.<br>2022 (13) | Cluster-RCT | Stratified versus<br>stepped psychological<br>care for depression | Clinician-level cluster randomization;<br>patients recruited after cluster allocation;<br>clinicians aware of allocation; unequal<br>numbers recruited across trial arms | Core domains and cluster<br>extension; recruitment<br>timing and potential<br>differential recruitment<br>(D1b) |
| Taneja et al.<br>2007 (4) | Crossover<br>RCT | Modafinil crossover<br>intervention | Two-period design; 4-day washout; order<br>effects assessed; carryover modelled | Core domains and<br>crossover extension;<br>adequate mitigation of<br>potential carryover (D6) |
| Holze et al.<br>2023 (5) | Crossover<br>RCT | LSD-assisted therapy<br>for anxiety | Randomized two-period crossover;<br>potentially persistent treatment effects;<br>acknowledged carryover; primary<br>interpretation emphasized first-period<br>results | Core domains and<br>crossover extension;<br>detection and<br>interpretation of<br>clinically important<br>carryover (D6) |

Development studies were purposively selected to represent contrasting methodological scenarios capable of challenging the RoB 2 decision framework. The set included individually randomized, cluster-randomized, and crossover trials and was used for calibration and assessment of core- and extension-domain decision logic rather than estimation of system performance.

**Supplementary Table S2.** Characteristics and methodological features of studies included in the post-freeze evaluation.

| **Study ID (year)** | **Trial design** | **Clinical context / intervention** | **Key methodological features represented** | **RoB 2 component represented** |
| --- | --- | --- | --- | --- |
| Blumberger et al. 2018 (14) | Parallel-group RCT | Neuromodulation intervention | Allocation concealment, missing outcome data, reporting considerations | Core domains |
| Mukherjee et al. 2023 (15) | Parallel-group RCT | Psychiatric intervention | Deviations from intended interventions, attrition handling | Core domains |
| Fröbert et al. 2013 (16) | Parallel-group RCT | Cardiovascular intervention | Randomization procedures, objective outcome measurement | Core domains |
| Ahmad et al. 2021 (17) | Parallel-group RCT | Clinical therapeutic intervention | Missing outcome data, selective-reporting considerations | Core domains |
| Baeken et al. 2013 (18) | Crossover RCT | Neuromodulation intervention | No formal washout, potential persistence of intervention effects, carryover considerations | Core domains and crossover extension |
| Sha et al. 2015 (19) | Crossover RCT | Pharmacodynamic intervention | 12–14-day washout, mixed-effects modelling, period and sequence effects | Core domains and crossover extension |
| Broughton et al. 1997 (20) | Crossover RCT | Modafinil for narcolepsy | Three-period crossover, assessment of carryover, analytical mitigation | Core domains and crossover extension |
| Garcia et al. 2020 (21) | Cluster-randomized trial | Cluster-level intervention | Participant recruitment after cluster randomization, potential differential recruitment | Core domains and cluster extension |
| Hayes et al. 2019 (22) | Cluster-randomized trial | Pragmatic cluster intervention | Recruitment procedures and timing relative to cluster randomization | Core domains and cluster extension |
| Cox et al. 2020 (23) | Cluster-randomized trial | Pragmatic cluster intervention | Post-randomization recruitment and recruitment-related considerations | Core domains and cluster extension |

**Note:** These 10 studies constituted the post-freeze evaluation dataset and were separate from the studies used for development and calibration. RoB2-Evaluator was frozen before assessment of the evaluation dataset, and no modifications to the instruction framework were permitted during subsequent temporal-stability or human-agreement analyses

**Supplementary Table S3.** Study-level temporal stability of RoB2-Evaluator across repeated assessments.

| **Study** | **Overall RoB, T1** | **Overall RoB, T2** | **Overall judgment concordant** | **Domain(s) with changed judgment\*** |
| --- | --- | --- | --- | --- |
| Blumberger et al., 2018 | Low | Low | Yes | None |
| Mukherjee et al., 2023 | Some concerns | Some concerns | Yes | D2, D3 |
| Fröbert et al., 2013 | Low | Low | Yes | None |
| Ahmad et al., 2021 | Some concerns | Some concerns | Yes | D2 |
| Baeken et al., 2013 | High | High | Yes | D3 |
| Sha et al., 2015 | Low | Some concerns | No | D1, D6 |
| Broughton et al., 1997 | High | High | Yes | D1 |
| Garcia et al., 2020 | High | High | Yes | D2 |
| Hayes et al., 2019 | Some concerns | Some concerns | Yes | None |
| Cox et al., 2020 | Some concerns | Some concerns | Yes | D2 |

*\* Domain changes indicate differences between T1 and T2 domain-level judgments. Domain-level changes did not necessarily alter the overall RoB judgment. D1b showed complete agreement across all applicable cluster-randomized trials.*

**Supplementary Table S4.** Temporal consistency of AI-generated methodological rationales and reason–decision alignment.

| **Evaluation** | **Classification / definition** | **n/N (%)** |
| --- | --- | --- |
| Reasoning consistency | Identical rationale | 31/53 (58.5) |
|  | Semantically equivalent rationale | 19/53 (35.8) |
|  | Different rationale | 3/53 (5.7) |
|  | Identical or semantically equivalent | 50/53 (94.3) |
| Reason–decision alignment | Rationale logically consistent with assigned RoB 2 judgment | 53/53 (100) |

Note: Reasoning consistency compared the methodological rationales generated at T1 and T2. Rationales were classified as **identical** when the substantive justification and wording were essentially unchanged; **semantically equivalent** when wording or emphasis differed but the same underlying methodological interpretation was expressed; or **different** when the assessments reflected a substantively different methodological rationale or interpretation. Overall reasoning consistency was defined as the proportion classified as identical or semantically equivalent. Reason–decision alignment assessed whether each generated rationale logically supported its corresponding RoB 2 judgment and should not be interpreted as evidence of judgment accuracy.

**Supplementary Table S5:**

| RoB 2 domain | Comparisons, n | Full alignment, n (%) | Partial alignment, n (%) | Conceptual disagreement, n (%) |
| --- | --- | --- | --- | --- |
| Domain 1 | 10 | 8 (80%) | 2 (20%) | 0 (0%) |
| Domain 2 | 10 | 9 (90%) | 1 (10%) | 0 (0%) |
| Domain 3 | 10 | 9 (90%) | 1 (10%) | 0 (0%) |
| Domain 4 | 10 | 10 (100%) | 0 (0%) | 0 (0%) |
| Domain 5 | 10 | 6 (60%) | 4 (40%) | 0 (0%) |
| Domain 1b<br>(cluster trials)<sup>†</sup> | 3 | 3 (100%) | 0 (0%) | 0 (0%) |
| Domain 6<br>(crossover trials)<sup>‡</sup> | 3 | 3 (100%) | 0 (0%) | 0 (0%) |
| **Overall** | **56** | **48 (85.7%)** | **8 (14.3%)** | **0 (0%)** |

Note: Full alignment indicates that RoB2-Evaluator and the human reviewers identified the same methodological concern and provided comparable justifications. Partial alignment indicates that both identified the same underlying source of bias but differed in the level of explanatory detail or emphasis. Conceptual disagreement indicates disagreement regarding the underlying methodological source of bias.
<sup>†</sup> Assessed only in cluster-randomized trials ( $n = 3$ ).
<sup>‡</sup> Assessed only in crossover trials ( $n = 3$ ).

**Supplementary Table S6.** Study-level conceptual alignment between RoB2-Evaluator and human consensus assessments.

| Study | D1 | D2 | D3 | D4 | D5 | D1b | D6 | Overall conceptual alignment |
| --- | --- | --- | --- | --- | --- | --- | --- | --- |
| Blumberger et al., 2018 | Full | Full | Full | Full | Full | — | — | Full |
| Mukherjee et al., 2023 | Partial | Full | Full | Full | Partial | — | — | Partial |
| Fröbert et al., 2013 | Full | Full | Full | Full | Full | — | — | Full |
| Ahmad et al., 2021 | Full | Full | Full | Full | Partial | — | — | Partial |
| Baeken et al., 2013 | Full | Full | Full | Full | Full | — | Full | Full |
| Sha et al., 2015 | Partial | Full | Full | Full | Full | — | Full | Partial |
| Broughton et al., 1997 | Full | Full | Full | Full | Full | — | Full | Full |
| Garcia et al., 2020 | Full | Partial | Full | Full | Partial | Full | — | Partial |
| Hayes et al., 2019 | Full | Full | Partial | Full | Full | Full | — | Partial |
| Cox et al., 2020 | Full | Full | Full | Full | Full | Full | — | Full |

**Full alignment** indicates that RoB2-Evaluator and the human consensus assessment identified the same underlying methodological concern and provided substantively comparable rationales. **Partial alignment** indicates identification of the same underlying methodological concern with differences in explanatory detail or emphasis. **Conceptual disagreement** indicates different interpretations of the underlying methodological issue. No conceptual disagreements were observed. Overall conceptual alignment was classified as Full when all applicable domains showed full alignment and Partial when at least one applicable domain showed partial alignment in the absence of conceptual disagreement. Conceptual alignment was evaluated independently of concordance in the resulting RoB 2 judgment. D1b was applicable only to cluster-randomized trials and D6 only to crossover trials; — indicates not applicable.

## Appendix B

### Supplementary Appendix B. RoB2-Evaluator Operating Instructions, Prompting Guide, and Output Templates

#### A1. Purpose and scope

RoB2-Evaluator was designed to support structured application of the Cochrane Risk of Bias 2 (RoB 2) framework to randomized controlled trials. The system used a fixed RoB 2–specific methodological knowledge base and a locked instruction framework to guide assessments through the applicable signalling questions and decision pathways.

RoB2-Evaluator was configured for:

- individually randomized parallel-group trials;
- cluster-randomized trials, including the recruitment domain (D1b); and
- crossover trials, including assessment of carryover effects (D6).

Unless otherwise specified, the default effect of interest was the **effect of assignment to intervention**, corresponding to an intention-to-treat–type estimand.

#### A2. Fixed methodological knowledge base

The Version 3 system used a fixed methodological knowledge base comprising:

1. *Cochrane Handbook for Systematic Reviews of Interventions*, Chapter 8;
2. Cochrane RoB 2 guidance for individually randomized parallel-group trials;
3. RoB 2 guidance for cluster-randomized trials; and
4. RoB 2 guidance for crossover trials.

Web browsing and external database access were not used during assessment. Trial-specific judgments were based on the supplied trial documentation together with the fixed methodological guidance available to the system.

#### A3. Trial documentation supplied for assessment

For each study, available trial documentation was supplied before initiation of the RoB 2 assessment, including:

- full-text primary publication;
- trial protocol, when available;
- statistical analysis plan, when available;
- trial-registry record, when available; and
- relevant supplementary material.

The system was instructed not to infer trial-specific facts that were unsupported by the supplied documentation.

#### A4. Core operating principles

The following principles were applied throughout assessment:

- Follow the applicable RoB 2 signalling questions and decision pathways.
- Distinguish absence of information from evidence of a methodological limitation.
- Do not apply undocumented numerical thresholds for missing outcome data or other domains.
- Do not automatically escalate D2 because a trial is open label or masking is absent.
- Do not infer allocation concealment from participant masking, identical treatment appearance, overencapsulation, or similar procedures unless concealment of the allocation sequence is supported by the trial documentation.
- Do not automatically escalate D5 solely because a protocol or statistical analysis plan is unavailable.
- Keep supporting evidence and methodological reasoning specific to the domain being assessed.
- Apply cluster-randomized and crossover extensions only when the corresponding trial design is present.
- Derive the overall RoB 2 judgment from the applicable domain-level judgments and RoB 2 decision rules rather than from an independent global impression.

These constraints were refined during development and calibration and incorporated into the frozen Version 3 framework.

#### A5. Structured prompting workflow

RoB2-Evaluator was operated using a standardized sequence of prompts. The workflow consisted of: (1) study-level assessment, (2) continuation to subsequent studies, (3) generation of the consolidated RoB 2 summary table, and (4) generation of the detailed master table.

Previously completed judgments were preserved during consolidation and were not reassessed or modified.

##### A5.1. Study-level assessment prompt

The following prompt initiated assessment of each RCT:

### RoB2-Evaluator Study Assessment Prompt

Assess the uploaded randomized controlled trial using the Cochrane Risk of Bias 2 (RoB 2) framework and the fixed methodological guidance available to RoB2-Evaluator.

#### 1. Determine the trial design

Identify whether the study is:

- an individually randomized parallel-group trial;
- a cluster-randomized trial; or
- a crossover trial.

Apply the corresponding RoB 2 framework:

- parallel-group trial: Domains 1–5;
- cluster-randomized trial: Domains 1–5 plus the recruitment domain (D1b), where applicable;
- crossover trial: Domains 1–5 plus the carryover domain (D6), where applicable.

#### 2. Define the effect of interest

Unless otherwise specified, assess the effect of assignment to intervention.

#### 3. Assess each applicable domain

For each domain:

- apply the relevant RoB 2 signalling questions and decision pathway;
- identify the trial information supporting the assessment;
- distinguish absence of information from evidence of a methodological limitation;
- provide a concise methodological rationale; and
- assign one of the following judgments: **Low risk**, **Some concerns**, or **High risk**.

Do not introduce unsupported assumptions, undocumented numerical thresholds, or methodological rules outside the applicable RoB 2 guidance. Keep evidence and reasoning specific to the domain being assessed.

#### 4. Determine the overall judgment

After completing all applicable domains, assign the overall RoB 2 judgment according to the applicable RoB 2 decision rules and provide a concise rationale.

#### 5. Output

Report:

- study identifier;
- trial design;
- effect of interest;
- each applicable domain judgment;
- supporting rationale for each domain; and
- overall RoB 2 judgment and rationale.

##### A5.2. Design-specific assessment instructions

###### Cluster-randomized trials

For cluster-randomized trials, additionally assess recruitment-related bias by considering:

- timing of participant identification and recruitment relative to cluster randomization;
- whether recruiters or individuals identifying participants knew the cluster allocation;
- whether knowledge of allocation could have influenced participant identification or recruitment;
- evidence of differential recruitment between intervention groups;
- baseline differences that may indicate recruitment-related selection; and
- whether any identified recruitment process could plausibly bias the intervention-effect estimate.

Post-randomization recruitment alone should not determine the D1b judgment. Recruitment timing should be interpreted together with knowledge of allocation, the potential for differential recruitment, and other relevant study information.

###### Crossover trials

For crossover trials, additionally assess potential carryover by considering:

- reversibility and expected persistence of the intervention effect;
- presence and adequacy of any washout period;
- treatment sequence and period effects;
- evidence of residual intervention effects;
- analytical approaches addressing period, sequence, or carryover effects; and
- whether carryover could plausibly bias the intervention-effect estimate.

The absence of a formal washout period should not automatically determine the D6 judgment. Washout should be interpreted in relation to the intervention, outcome, trial design, and analytical approach.

##### A5.3. Sequential study prompt

After completion of one study, subsequent trials were assessed using the same locked framework and output structure.

###### Next-Study Prompt

Assess the newly uploaded trial using the same RoB2-Evaluator framework, methodological guidance, effect-of-interest convention, domain definitions, decision rules, and output structure used for the preceding assessments.

Treat the study as a new RoB 2 assessment. Base all trial-specific judgments only on the documentation supplied for the current study.

Do not modify previously completed study assessments.

This procedure was repeated until all studies in the assessment set had been completed.

#### A6. Standard study-level output structure

Each completed study assessment generated the following structured output:

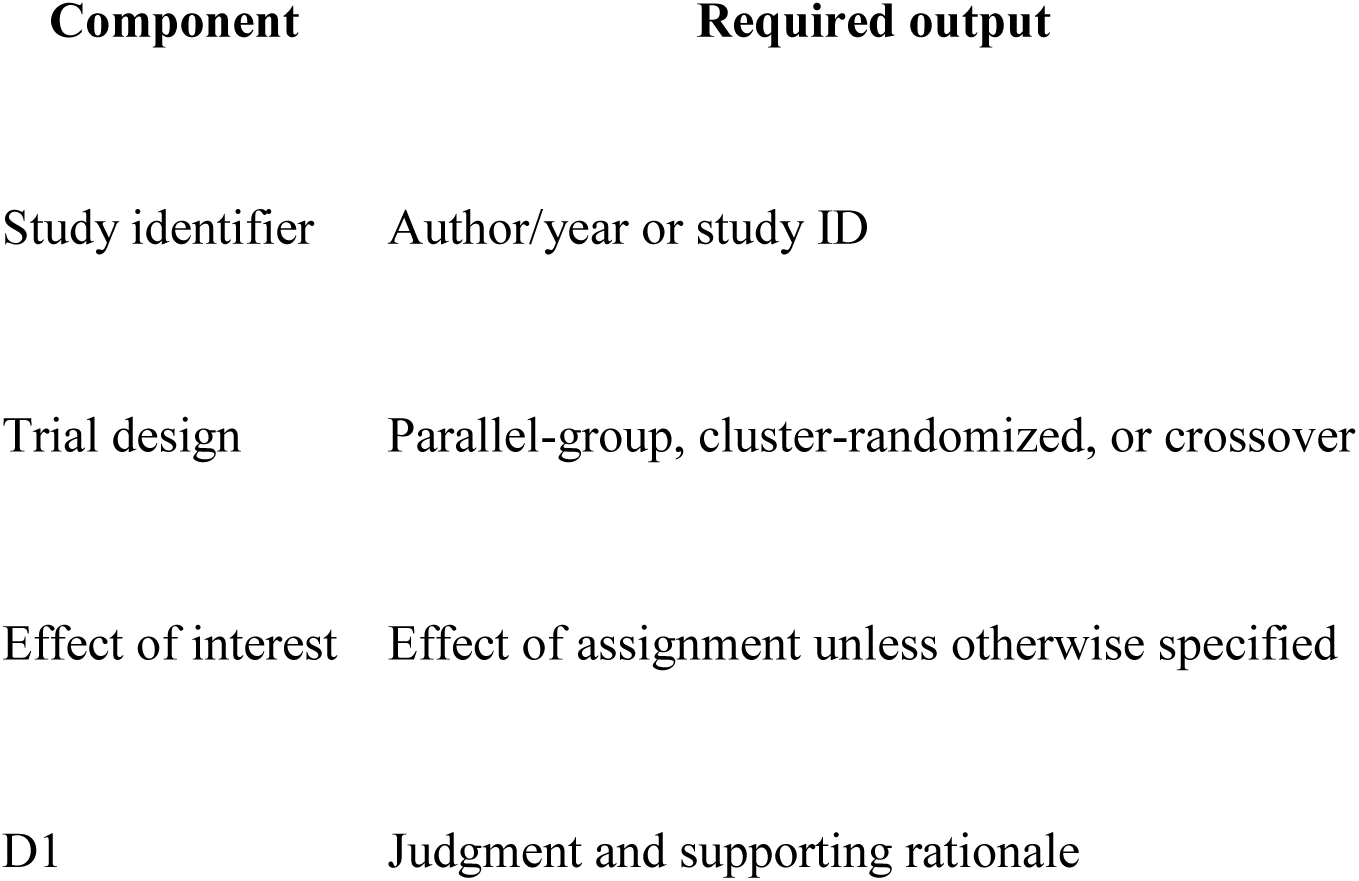
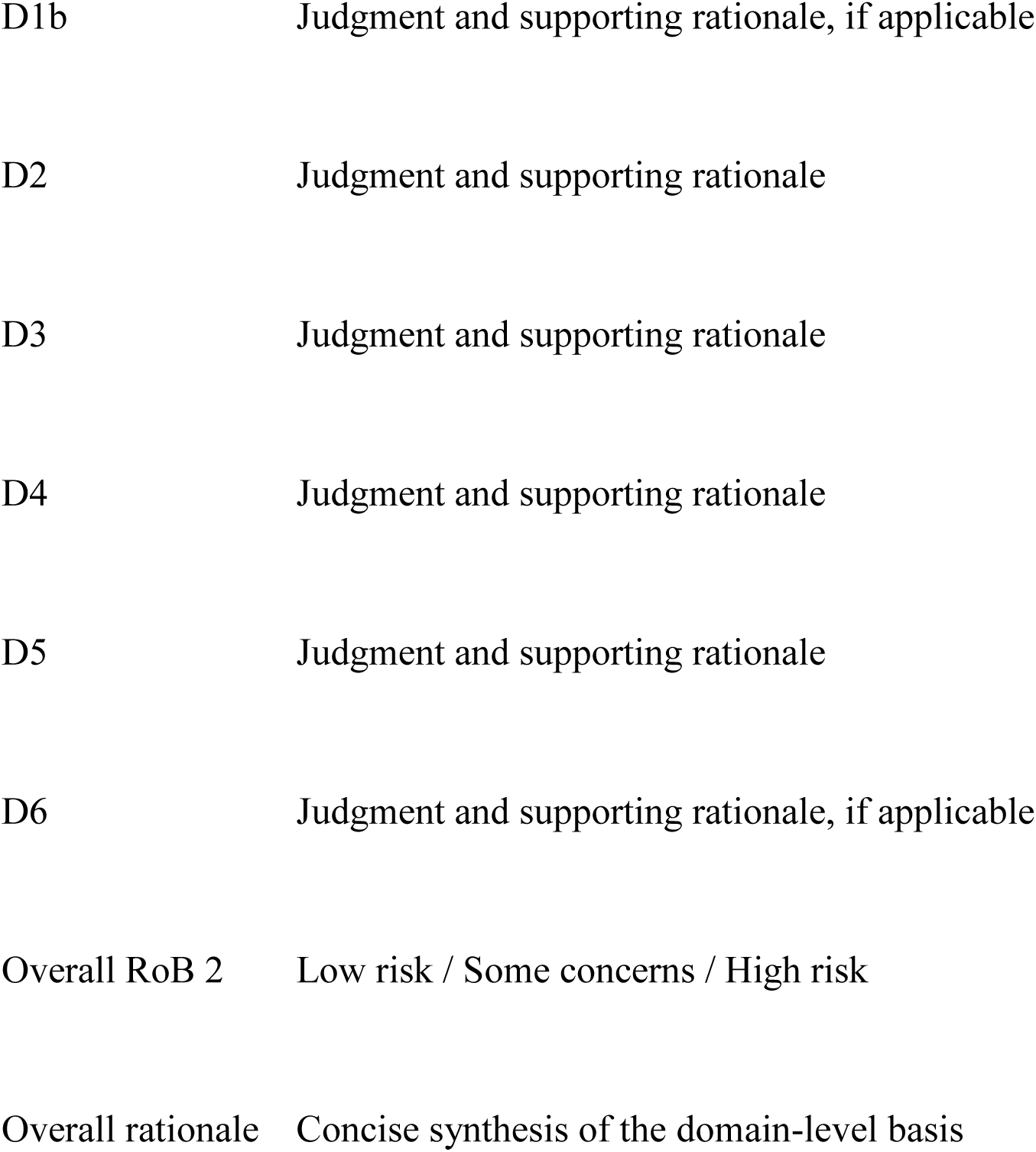

#### A7. Consolidated RoB 2 summary-table prompt

After all individual study assessments were completed, a standardized prompt was used to generate the study-level summary table.

##### RoB2 Summary Table Prompt

Compile all completed RoB2-Evaluator assessments into a single RoB 2 summary table.

Include **one row per study** and the following columns:

- Study ID;
- D1;
- D1b, where applicable;
- D2;
- D3;
- D4;
- D5;
- D6, where applicable; and
- Overall RoB 2 judgment.

Preserve the judgments assigned in the original study-level assessments.

Do not reassess, reinterpret, change, harmonize, or infer new judgments while constructing the table.

Use **—** where a design-specific domain is not applicable.

##### Required summary-table structure

| Study | D1 | D1b | D2 | D3 | D4 | D5 | D6 | Overall |
| --- | --- | --- | --- | --- | --- | --- | --- | --- |
| Study 1 | Judgment | — / Judgment | Judgment | Judgment | Judgment | Judgment | — / Judgment | Judgment |

The resulting table provided the structured data used for subsequent agreement analyses and graphical reporting.

#### A8. RoB 2 master-table prompt

A second standardized prompt generated a detailed table containing both judgments and their supporting methodological rationales.

##### RoB2 Master Table Prompt

Compile all completed RoB2-Evaluator study assessments into a detailed master table.

For each study and each applicable RoB 2 domain, report:

- domain judgment;
- supporting methodological rationale; and
- overall RoB 2 judgment.

Preserve the content of the completed study assessments.

Do not reassess, reinterpret, revise, or modify previously assigned judgments or rationales when compiling the table.

Use **—** for design-specific domains that are not applicable.

##### Required master-table structure

| Study | Domain | RoB 2 judgment | Supporting methodological rationale | Overall RoB 2 |
| --- | --- | --- | --- | --- |
| Study 1 | D1 | Low / Some concerns / High | Study-specific rationale | Overall judgment |
| Study 1 | D2 | Low / Some concerns / High | Study-specific rationale | Overall judgment |

The master table served as the source for evaluation of methodological-rationale consistency and AI–human conceptual alignment.

#### A9. Consolidation safeguards

The following constraints were applied when generating summary and master tables:

- completed study-level assessments were treated as final;
- table-generation prompts did not initiate a new RoB 2 assessment;
- judgments were transferred without reinterpretation or harmonization;
- rationales were preserved without substantive modification;
- unavailable or non-applicable design-specific domains were not inferred; and
- generation of consolidated tables did not permit modification of the frozen assessment framework.

#### A10. Version control

| **Version** | **Development stage** | **Status** |
| --- | --- | --- |
| Version 1 | Initial GPT-5.2 implementation with fixed RoB 2 methodological knowledge base and locked instruction framework | Development |
| Version 2 | Core-domain calibration completed | Frozen core framework |
| Version 3 | Cluster-randomized and crossover extension calibration completed | Final frozen evaluation framework |

No modifications to the instruction framework were permitted after Version 3 was frozen for the post-freeze evaluation.

#### A11. Reproducibility considerations

RoB2-Evaluator uses a proprietary underlying large language model and execution environment that may change independently of the locked instruction framework. Reproduction of the assessment procedure therefore requires preservation of the system instructions, methodological knowledge files, standardized prompts, model identifier, system version, and assessment date.

Version freezing in this study refers to the RoB2-Evaluator instruction and methodological knowledge architecture and does not imply control over subsequent changes to the underlying model or execution environment.

